# Genetic and Shared Environmental Influences on Cancer Risk and Cross-Cancer Associations in Nordic Twins

**DOI:** 10.64898/2026.06.18.26355861

**Authors:** Jennifer R. Harris, Signe B. Clemmensen, Hans-Olov Adami, Lorelei Mucci, Jaakko Kaprio, Jacob v.B. Hjelmborg

## Abstract

The relative contributions of genetic and shared environmental influences to cancer risk and cross-cancer associations remain poorly understood. We analyzed data from 222,530 same-sex twins from Denmark, Finland, Norway, and Sweden in the Nordic Twin Study of Cancer, including 43,060 incident cancers over a median follow-up of 41.6 years. Using a target trial framework, biometric modeling, and competing-risk adjustment, we estimated familial risk, heritability, and shared environmental contributions across 35 cancer sites. Lifetime cancer risk was 36.5%, increasing to 51.4% in monozygotic (MZ) twins and 45.3% in dizygotic (DZ) twins with an affected co-twin. Overall cancer risk was explained by heritable (28%) and shared environmental (40%) influences. Heritability was highest for prostate (42%), non-melanoma skin (24%), and breast (18%) cancers. Cross-cancer analyses revealed extensive overlap in the genetic and shared environmental factors across sites, consistent with widespread pleiotropy and shared environmental susceptibility. Prostate cancer exhibited the strongest genetic overlap with rectum/anus (12%) and kidney (11%) cancers, whereas co-shared environmental influences were most pronounced for breast-lung (11%), prostate-bladder (11%), and prostate-lung (12%) cancers. These findings show pervasive genetic overlap across cancers at different sites and emphasize the importance of incorporating familial shared environmental exposures into cancer risk prediction and prevention strategies.

## INTRODUCTION

Elucidating the genetic risk of cancer is critical to understanding its etiology and guiding risk stratification. Clustering of cancers in families, whether for an individual cancer or multiple cancers, can reflect shared genes and shared exposures to cancer risk factors. While cancer predisposition syndromes with high penetrance pathogenic variants account for a portion of cancer familial risk, a considerable burden remains unexplained. Twin^1^, family^2^, and germline genetic^3–6^ studies have provided estimates of heritable variation underlying cancer risk at multiple sites. Genome-wide association (GWA) studies for multiple cancers have identified genetic risk loci underlying this heritable risk.

A systematic review provided early evidence that many cancer-associated genes and loci exhibit pleiotropic effects across multiple cancers^7^. Subsequent large-scale pan-cancer GWAs and biobank-based studies support this observation, demonstrating extensive sharing of genetic susceptibility across cancers. Several studies report genetic correlations or associations between sets of cancers^3–6,8–12^. Patterns of cross-cancer pleiotropy are also emerging from the analysis of associations between the polygenic risk score for certain cancers and specific cancer outcomes^8,13^. This body of research reveals that pleiotropy is widespread^5,7,9,10,14–21^, pervasive for hormone-related cancers, and suggests that lead genetic variants can confer differential effects depending upon the type of cancer^22^. Additionally, investigations have begun to identify the specific pleiotropic variants, loci, and pathways involved^4,5,8,9,12,14–21,23^. While environmental factors also influence cancer risk at multiple sites, their heritable variation^24–26^ can make it difficult to distinguish these effects from genetic pleiotropy among cancers.

Whereas germline studies of pleiotropy rely on identifying genetic variants shared across cancers, the twin classical design exploits the differential degree of genotype sharing between monozygotic (MZ) and dizygotic (DZ) twins to estimate coheritability. MZ twins are genetically identical at the sequence level, while members of DZ pairs share, on average, half of their segregating genes. Thus, the within-pair recurrence risk among MZ pairs reflects the risk for developing two different cancers associated with the same underlying genetic predisposition. Because the twin design captures both additive and non-additive genetic influences, sufficiently powered twin studies may yield estimates of coheritable factors that exceed those derived from GWA-based additive effects of currently identified cancer genetic risk variants. Moreover, the twin design helps determine the contribution of shared familial environments to the cross-cancer associations.

The largest published twin study of familial risk for cancers revealed that cancer concordance among twins often manifests across, rather than within, cancer sites^1^. If one twin had cancer, their co-twin was at an increased risk of developing cancer, but most often at a different anatomical site. Specifically, among twin pairs in which both developed cancer, 62 percent of the MZ pairs and 74% of DZ were discordant for the cancer site.

In this study, we estimate the genetic and environmental sources of covariation underlying cross-cancer occurrences in twins, leveraging updated data from the Nordic Twin Study of Cancer (NorTwinCan)^27^. Our analysis encompasses data from 222,530 same-sexed twins, 43,060 incident cancer cases spanning 35 cancer sites, and a median follow-up of more than 41 years. We provide new estimates of familial effects and heritability and, for the first time, report relative recurrence risks and coheritability across cancers. To facilitate comparisons with other studies, we calculate heritability using two approaches suited for censored time-to-event data in twins: the risk-scale model, which accounts for competing risk of death^28^, and the classical liability threshold model. The results aim to shed light on the underlying factors driving the observed cross-cancer associations.

## Results

Characteristics of the twin cohort are provided in **Table 1**. 40% of the twins were MZ and 52% were female. During a median follow-up of 41.6 years, 43,060 incident cancers were diagnosed; 69,063 of twins died of any cause, and 2,267 emigrated.

**Table 1.** Characteristics of the NorTwinCan cohort of 222,530 individual twins from same-sex pairs with follow-up for cancer incidence, 1943 to 2016.

| **Country** | **Denmark** | **Finland** | **Norway** | **Sweden** | **Total** |
| --- | --- | --- | --- | --- | --- |
| **Birth Cohort** | 1870-2004 | 1875-1957 | 1915-1991 | 1886-2008 |  |
| **N individual twins** | 70,345 | 26,957 | 28,984 | 96,244 | **222,530** |
| **N (%) MZ twins** | 25,733 (37) | 8,368 (31) | 13,100 (45) | 42,042 (44) | **89,243 (40)** |
| **N (%) female twins** | 34,640 (49) | 13,480 (50) | 15,546 (54) | 51,124 (53) | **114,790 (52)** |
| **First date follow-up** | Jan 1943 | Feb 1974 | April 1956 | Jan 1958 |  |
| **End of follow-up** | Dec 2016 | Dec 2016 | Dec 2015 | Dec 2015 |  |
| **Median years of follow-up (IQR)** | 47.7<br>(30.0-61.8) | 41.3<br>(28.9-41.3) | 56.2<br>(41.5-59.9) | 40.5<br>(22.3-58.2) | **41.6**<br>**(27.8-58.2)** |
| **Median entry age (IQR)** | 0.0 (0.0-13.8) | 32.3<br>(24.4-45.6) | 4.9<br>(0.0-18.3) | 3.5<br>(0.0-28.0) | **2.2**<br>**(0.0-28.2)** |
| **N incident cancers** | 13,067 | 6,002 | 5,367 | 18,624 | **43,060** |

### Site-specific findings

The overall lifetime risk of any cancer by age 100 years – after accounting for competing causes of death – was 36.5% (**Table 2**). Among the MZ pairs, 14,162 twins were diagnosed with cancer, including 2,502 MZ pairs where both twins had cancer (5,004 individuals) and 9,158 discordant pairs where only one twin was diagnosed with cancer. Among the DZ pairs, 23,990 twins were diagnosed with cancer, including 3,560 DZ pairs where both twins had cancer (7120 individuals), and 16,870 discordant pairs.

**Table 2.**
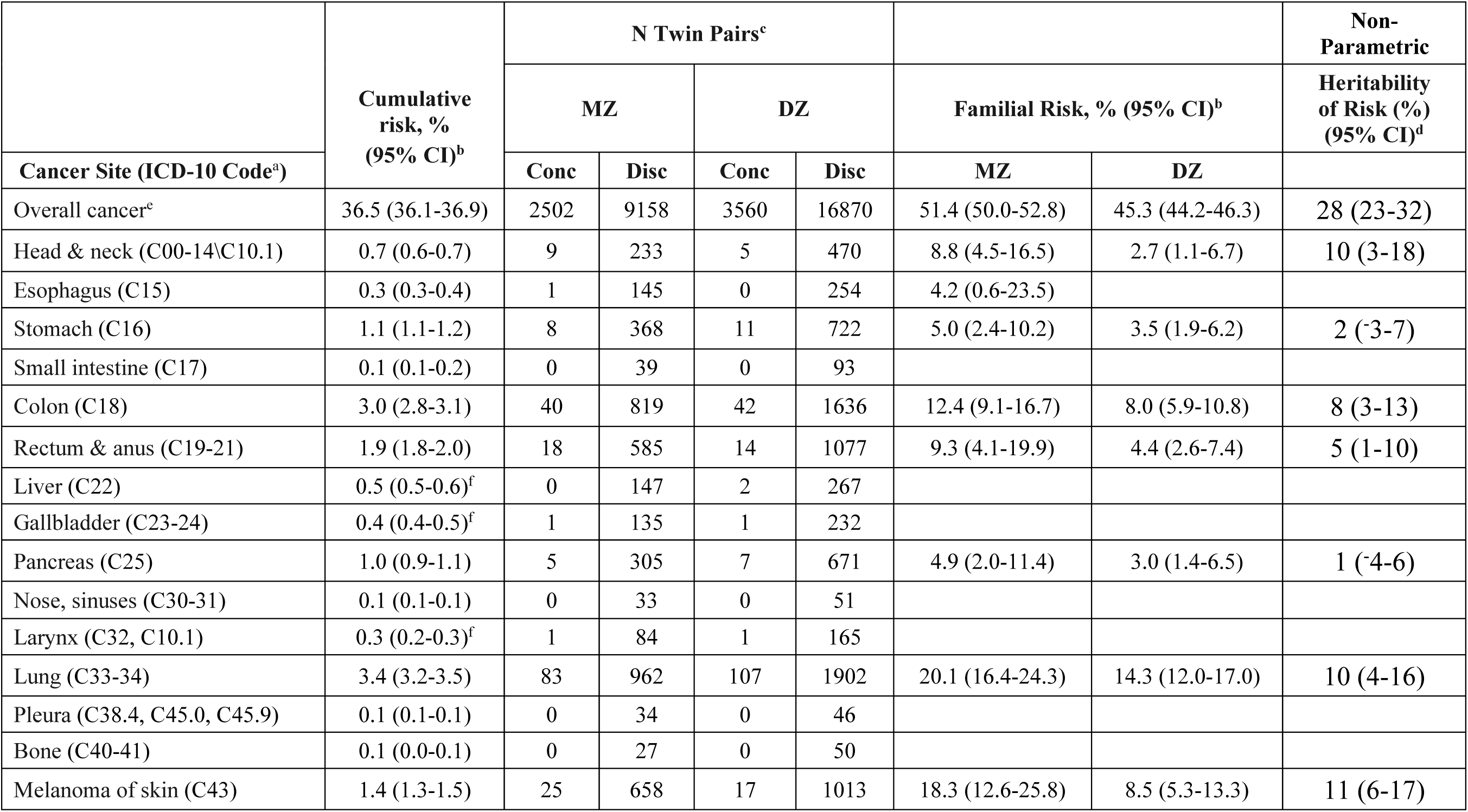
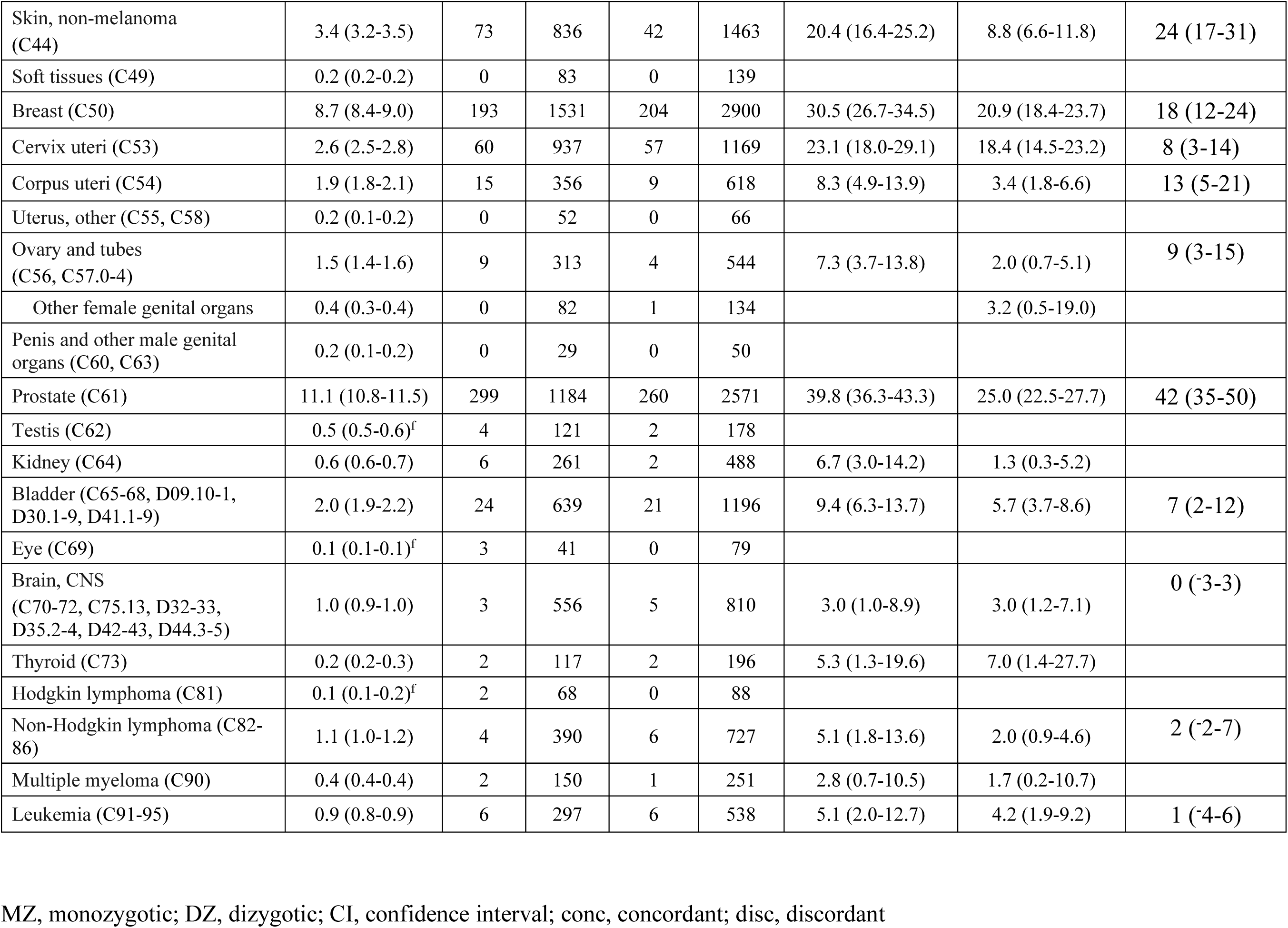
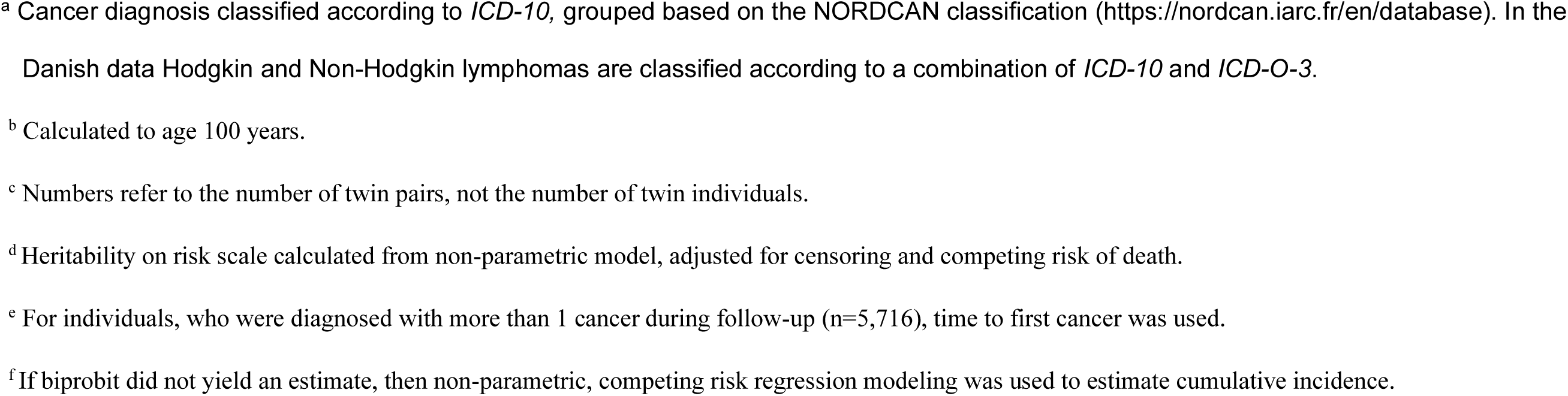
Cumulative and familial risk of cancer and heritability on the risk scale for selected malignancies among MZ and DZ twins adjusted for competing risk of death.

| Cancer Site (ICD-10 Code<sup>a</sup>) | Cumulative risk, % (95% CI)<sup>b</sup> | N Twin Pairs<sup>c</sup> |  |  |  |  |  | Non-Parametric |
| --- | --- | --- | --- | --- | --- | --- | --- | --- |
|  |  | MZ |  | DZ |  | Familial Risk, % (95% CI)<sup>b</sup> |  | Heritability of Risk (%) (95% CI)<sup>d</sup> |
|  |  | Conc | Disc | Conc | Disc | MZ | DZ |  |
| Overall cancer<sup>c</sup> | 36.5 (36.1-36.9) | 2502 | 9158 | 3560 | 16870 | 51.4 (50.0-52.8) | 45.3 (44.2-46.3) | 28 (23-32) |
| Head & neck (C00-14\C10.1) | 0.7 (0.6-0.7) | 9 | 233 | 5 | 470 | 8.8 (4.5-16.5) | 2.7 (1.1-6.7) | 10 (3-18) |
| Esophagus (C15) | 0.3 (0.3-0.4) | 1 | 145 | 0 | 254 | 4.2 (0.6-23.5) |  |  |
| Stomach (C16) | 1.1 (1.1-1.2) | 8 | 368 | 11 | 722 | 5.0 (2.4-10.2) | 3.5 (1.9-6.2) | 2 (-3-7) |
| Small intestine (C17) | 0.1 (0.1-0.2) | 0 | 39 | 0 | 93 |  |  |  |
| Colon (C18) | 3.0 (2.8-3.1) | 40 | 819 | 42 | 1636 | 12.4 (9.1-16.7) | 8.0 (5.9-10.8) | 8 (3-13) |
| Rectum & anus (C19-21) | 1.9 (1.8-2.0) | 18 | 585 | 14 | 1077 | 9.3 (4.1-19.9) | 4.4 (2.6-7.4) | 5 (1-10) |
| Liver (C22) | 0.5 (0.5-0.6)<sup>f</sup> | 0 | 147 | 2 | 267 |  |  |  |
| Gallbladder (C23-24) | 0.4 (0.4-0.5)<sup>f</sup> | 1 | 135 | 1 | 232 |  |  |  |
| Pancreas (C25) | 1.0 (0.9-1.1) | 5 | 305 | 7 | 671 | 4.9 (2.0-11.4) | 3.0 (1.4-6.5) | 1 (-4-6) |
| Nose, sinuses (C30-31) | 0.1 (0.1-0.1) | 0 | 33 | 0 | 51 |  |  |  |
| Larynx (C32, C10.1) | 0.3 (0.2-0.3)<sup>f</sup> | 1 | 84 | 1 | 165 |  |  |  |
| Lung (C33-34) | 3.4 (3.2-3.5) | 83 | 962 | 107 | 1902 | 20.1 (16.4-24.3) | 14.3 (12.0-17.0) | 10 (4-16) |
| Pleura (C38.4, C45.0, C45.9) | 0.1 (0.1-0.1) | 0 | 34 | 0 | 46 |  |  |  |
| Bone (C40-41) | 0.1 (0.0-0.1) | 0 | 27 | 0 | 50 |  |  |  |
| Melanoma of skin (C43) | 1.4 (1.3-1.5) | 25 | 658 | 17 | 1013 | 18.3 (12.6-25.8) | 8.5 (5.3-13.3) | 11 (6-17) |
| Skin, non-melanoma (C44) | 3.4 (3.2-3.5) | 73 | 836 | 42 | 1463 | 20.4 (16.4-25.2) | 8.8 (6.6-11.8) | 24 (17-31) |
| Soft tissues (C49) | 0.2 (0.2-0.2) | 0 | 83 | 0 | 139 |  |  |  |
| Breast (C50) | 8.7 (8.4-9.0) | 193 | 1531 | 204 | 2900 | 30.5 (26.7-34.5) | 20.9 (18.4-23.7) | 18 (12-24) |
| Cervix uteri (C53) | 2.6 (2.5-2.8) | 60 | 937 | 57 | 1169 | 23.1 (18.0-29.1) | 18.4 (14.5-23.2) | 8 (3-14) |
| Corpus uteri (C54) | 1.9 (1.8-2.1) | 15 | 356 | 9 | 618 | 8.3 (4.9-13.9) | 3.4 (1.8-6.6) | 13 (5-21) |
| Uterus, other (C55, C58) | 0.2 (0.1-0.2) | 0 | 52 | 0 | 66 |  |  |  |
| Ovary and tubes (C56, C57.0-4) | 1.5 (1.4-1.6) | 9 | 313 | 4 | 544 | 7.3 (3.7-13.8) | 2.0 (0.7-5.1) | 9 (3-15) |
| Other female genital organs | 0.4 (0.3-0.4) | 0 | 82 | 1 | 134 |  | 3.2 (0.5-19.0) |  |
| Penis and other male genital organs (C60, C63) | 0.2 (0.1-0.2) | 0 | 29 | 0 | 50 |  |  |  |
| Prostate (C61) | 11.1 (10.8-11.5) | 299 | 1184 | 260 | 2571 | 39.8 (36.3-43.3) | 25.0 (22.5-27.7) | 42 (35-50) |
| Testis (C62) | 0.5 (0.5-0.6)<sup>f</sup> | 4 | 121 | 2 | 178 |  |  |  |
| Kidney (C64) | 0.6 (0.6-0.7) | 6 | 261 | 2 | 488 | 6.7 (3.0-14.2) | 1.3 (0.3-5.2) |  |
| Bladder (C65-68, D09.10-1, D30.1-9, D41.1-9) | 2.0 (1.9-2.2) | 24 | 639 | 21 | 1196 | 9.4 (6.3-13.7) | 5.7 (3.7-8.6) | 7 (2-12) |
| Eye (C69) | 0.1 (0.1-0.1)<sup>f</sup> | 3 | 41 | 0 | 79 |  |  |  |
| Brain, CNS (C70-72, C75.13, D32-33, D35.2-4, D42-43, D44.3-5) | 1.0 (0.9-1.0) | 3 | 556 | 5 | 810 | 3.0 (1.0-8.9) | 3.0 (1.2-7.1) | 0 (-3-3) |
| Thyroid (C73) | 0.2 (0.2-0.3) | 2 | 117 | 2 | 196 | 5.3 (1.3-19.6) | 7.0 (1.4-27.7) |  |
| Hodgkin lymphoma (C81) | 0.1 (0.1-0.2)<sup>f</sup> | 2 | 68 | 0 | 88 |  |  |  |
| Non-Hodgkin lymphoma (C82-86) | 1.1 (1.0-1.2) | 4 | 390 | 6 | 727 | 5.1 (1.8-13.6) | 2.0 (0.9-4.6) | 2 (-2-7) |
| Multiple myeloma (C90) | 0.4 (0.4-0.4) | 2 | 150 | 1 | 251 | 2.8 (0.7-10.5) | 1.7 (0.2-10.7) |  |
| Leukemia (C91-95) | 0.9 (0.8-0.9) | 6 | 297 | 6 | 538 | 5.1 (2.0-12.7) | 4.2 (1.9-9.2) | 1 (-4-6) |

MZ, monozygotic; DZ, dizygotic; CI, confidence interval; conc, concordant; disc, discordant
<sup>a</sup> Cancer diagnosis classified according to *ICD-10*, grouped based on the NORDCAN classification (https://nordcan.iarc.fr/en/database). In the Danish data Hodgkin and Non-Hodgkin lymphomas are classified according to a combination of *ICD-10* and *ICD-O-3*.
<sup>b</sup> Calculated to age 100 years.
<sup>c</sup> Numbers refer to the number of twin pairs, not the number of twin individuals.
<sup>d</sup> Heritability on risk scale calculated from non-parametric model, adjusted for censoring and competing risk of death.
<sup>e</sup> For individuals, who were diagnosed with more than 1 cancer during follow-up (n=5,716), time to first cancer was used.
<sup>f</sup> If biprobit did not yield an estimate, then non-parametric, competing risk regression modeling was used to estimate cumulative incidence.

The lifetime risk for developing any cancer in twins whose co-twin also had a cancer diagnosis was 45.3% for DZ and 51.4% for MZ twins. Compared to the risk in the overall cohort, the risk was 9 percent higher among DZ twins and 15 percent higher among MZ twins if their co-twin also has a cancer diagnosis.

There was an excess familial risk for 21 of the site-specific cancers (**Table 2**). The estimates were greater among the MZ than the DZ pairs for nearly all the cancers, and these zygosity differences were statistically significant for overall cancer and for the more common cancers, including non-melanoma skin, breast, and prostate cancer. After correcting for the competing risk of death, the heritability for overall cancer risk was 28%. The cancer sites showing the highest heritability of risk were prostate (42%), non-melanoma skin (24%), breast (18%), and corpus uteri (13%) (**Table 2**).

Tetrachoric correlations, heritability of liability estimates, and shared family environmental effects derived from parametric models for censored time-to-event data are displayed in **Table 3** for cancer sites with estimable parameters. The expanded dataset enabled estimation of heritability using a parametric liability model for seven cancers that could not be estimated in the earlier NorTwinCan dataset^1^, including cancers of the pancreas, cervix uteri, thyroid, liver, non-Hodgkin lymphoma, Hodgkin lymphoma, and multiple myeloma.

**Table 3.**
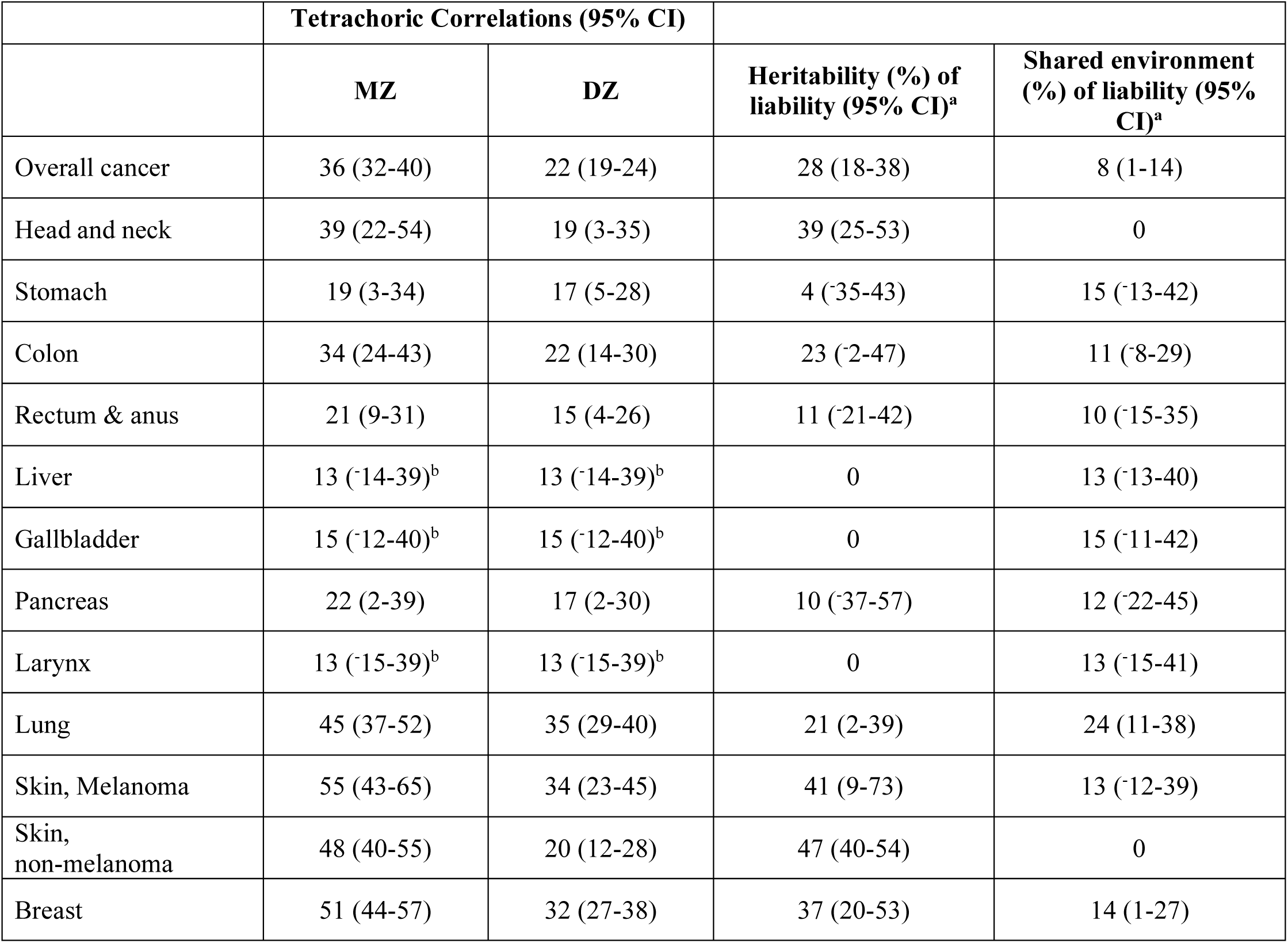
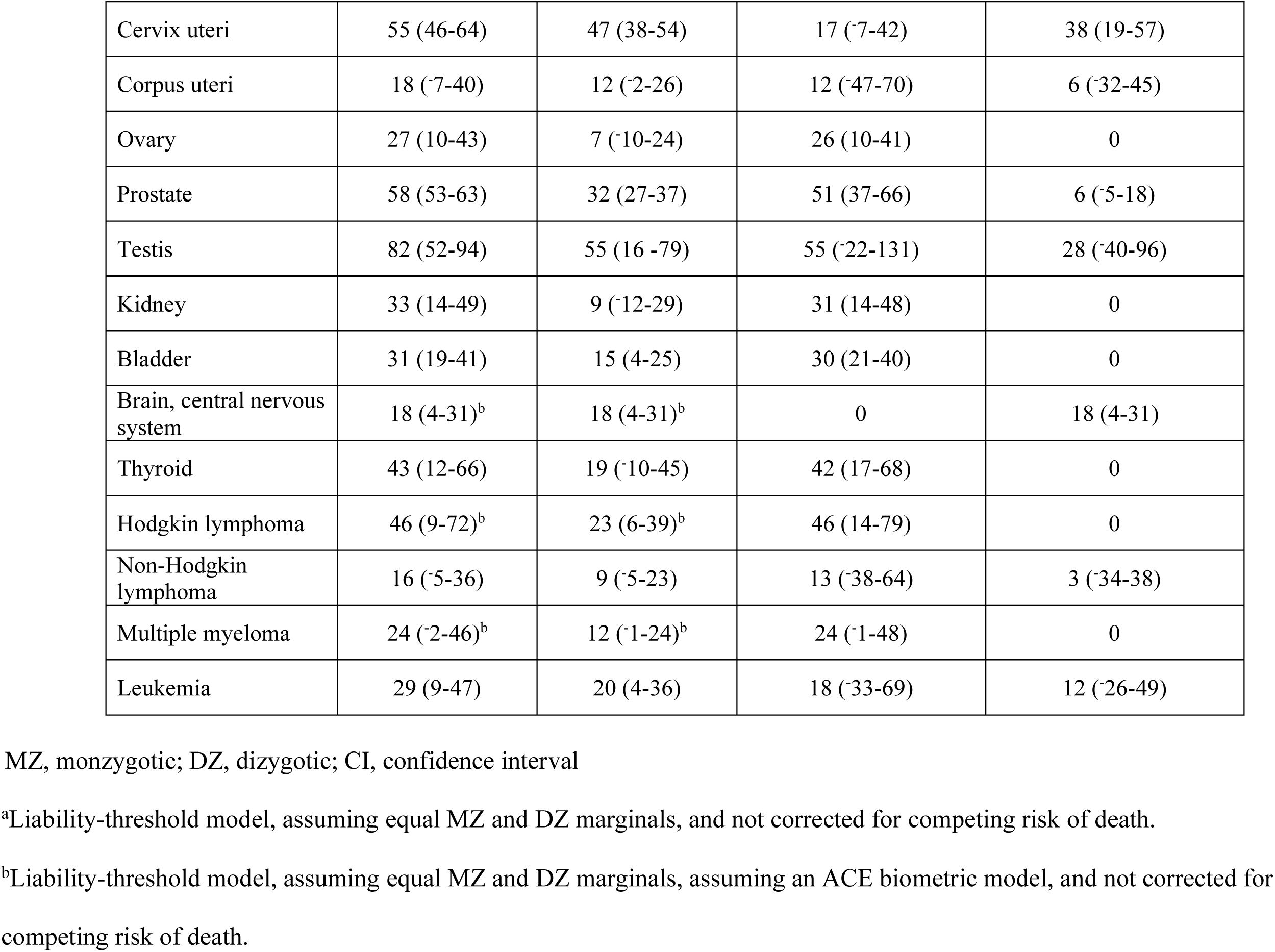
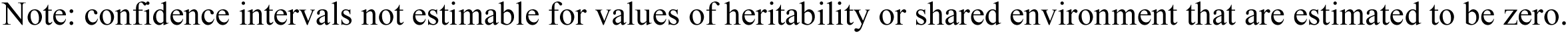
Tetrachoric correlations and estimates of heritability and shared environmental effects for select cancers based on censored time-to-event data from the parametric liability threshold model.

|  | **Tetrachoric Correlations (95% CI)** |  |  |  |
| --- | --- | --- | --- | --- |
|  | **MZ** | **DZ** | **Heritability (%) of liability (95% CI)<sup>a</sup>** | **Shared environment (%) of liability (95% CI)<sup>a</sup>** |
| Overall cancer | 36 (32-40) | 22 (19-24) | 28 (18-38) | 8 (1-14) |
| Head and neck | 39 (22-54) | 19 (3-35) | 39 (25-53) | 0 |
| Stomach | 19 (3-34) | 17 (5-28) | 4 (-35-43) | 15 (-13-42) |
| Colon | 34 (24-43) | 22 (14-30) | 23 (-2-47) | 11 (-8-29) |
| Rectum & anus | 21 (9-31) | 15 (4-26) | 11 (-21-42) | 10 (-15-35) |
| Liver | 13 (-14-39)<sup>b</sup> | 13 (-14-39)<sup>b</sup> | 0 | 13 (-13-40) |
| Gallbladder | 15 (-12-40)<sup>b</sup> | 15 (-12-40)<sup>b</sup> | 0 | 15 (-11-42) |
| Pancreas | 22 (2-39) | 17 (2-30) | 10 (-37-57) | 12 (-22-45) |
| Larynx | 13 (-15-39)<sup>b</sup> | 13 (-15-39)<sup>b</sup> | 0 | 13 (-15-41) |
| Lung | 45 (37-52) | 35 (29-40) | 21 (2-39) | 24 (11-38) |
| Skin, Melanoma | 55 (43-65) | 34 (23-45) | 41 (9-73) | 13 (-12-39) |
| Skin, non-melanoma | 48 (40-55) | 20 (12-28) | 47 (40-54) | 0 |
| Breast | 51 (44-57) | 32 (27-38) | 37 (20-53) | 14 (1-27) |
| Cervix uteri | 55 (46-64) | 47 (38-54) | 17 (-7-42) | 38 (19-57) |
| Corpus uteri | 18 (-7-40) | 12 (-2-26) | 12 (-47-70) | 6 (-32-45) |
| Ovary | 27 (10-43) | 7 (-10-24) | 26 (10-41) | 0 |
| Prostate | 58 (53-63) | 32 (27-37) | 51 (37-66) | 6 (-5-18) |
| Testis | 82 (52-94) | 55 (16 -79) | 55 (-22-131) | 28 (-40-96) |
| Kidney | 33 (14-49) | 9 (-12-29) | 31 (14-48) | 0 |
| Bladder | 31 (19-41) | 15 (4-25) | 30 (21-40) | 0 |
| Brain, central nervous system | 18 (4-31)<sup>b</sup> | 18 (4-31)<sup>b</sup> | 0 | 18 (4-31) |
| Thyroid | 43 (12-66) | 19 (-10-45) | 42 (17-68) | 0 |
| Hodgkin lymphoma | 46 (9-72)<sup>b</sup> | 23 (6-39)<sup>b</sup> | 46 (14-79) | 0 |
| Non-Hodgkin lymphoma | 16 (-5-36) | 9 (-5-23) | 13 (-38-64) | 3 (-34-38) |
| Multiple myeloma | 24 (-2-46)<sup>b</sup> | 12 (-1-24)<sup>b</sup> | 24 (-1-48) | 0 |
| Leukemia | 29 (9-47) | 20 (4-36) | 18 (-33-69) | 12 (-26-49) |

MZ, monzygotic; DZ, dizygotic; CI, confidence interval
<sup>a</sup>Liability-threshold model, assuming equal MZ and DZ marginals, and not corrected for competing risk of death.
<sup>b</sup>Liability-threshold model, assuming equal MZ and DZ marginals, assuming an ACE biometric model, and not corrected for competing risk of death.
Note: confidence intervals not estimable for values of heritability or shared environment that are estimated to be zero.

The parametric model (**Table 3**) consistently yields higher heritability estimates and wider confidence intervals than the non-parametric model (**Table 2**). However, when values were transformed to a uniform (risk) scale, the heritability estimates from these two models converged considerably (**Figure S1**). The parametric modeling revealed that shared environmental factors contributed to variance in cancer risk at many cancer sites **(Table 3)**. The updated analyses also yielded new estimates of shared environmental effects for 17 additional cancers beyond our previous report^1^ and provided refined estimates for shared environmental effects for cancers of the lung (0.24), breast (0.14), cervix uteri (0.38), brain (0.18), and for overall cancer (0.08).

### Cross-cancer analyses

Most of the cancer-concordant pairs were discordant for the anatomic cancer site. Overall, 62% of the cancer concordant MZ pairs and 74% of the cancer concordant DZ pairs developed different cancers. Among the MZ pairs where both developed cancer, 31% (788 pairs) were concordant for the specific cancer type when based on the first diagnosed cancer in each twin. This concordance increased to 38% (960 pairs) when based on any cancer diagnoses in each twin. The corresponding values for cancer concordant DZ pairs revealed that 22% (770 pairs) of the pairs developed cancer at the same site when based on their first diagnosis, and 26% (916 pairs) were concordant for cancer at the same site when all diagnoses were accounted for regardless of chronology. **Table S1** presents the cross-tabulations showing the pairwise distribution of the number of cancer diagnoses in the twins, stratified by zygosity.

**Table 4** presents the lifetime relative recurrence risk (RRR) for cancer co-occurrences within MZ and DZ pairs across 32 cancer sites, ordered along the diagonal by the MZ RRR. The clustering of elevated RRR values in the lower right portion of the table indicates that familial factors contribute importantly to many cross-cancer associations. Among the MZ pairs, the RRR were more than double the DZ value for numerous cross-cancer pairs. Although the RRRs are generally lower among DZ pairs, many cross-cancer associations exceeded 1.0, consistent with a contribution of shared familial factors.

**Table 4.**
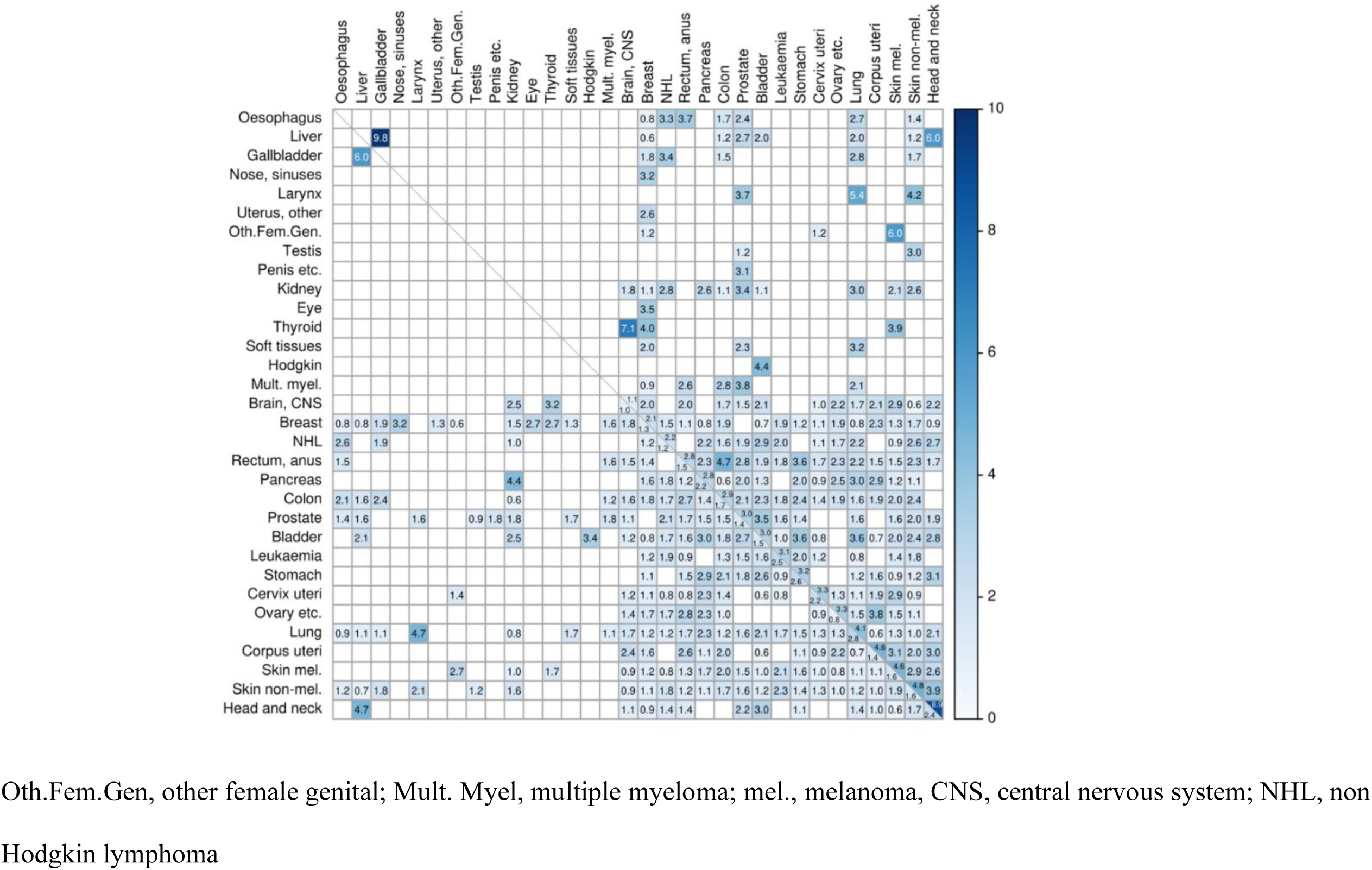
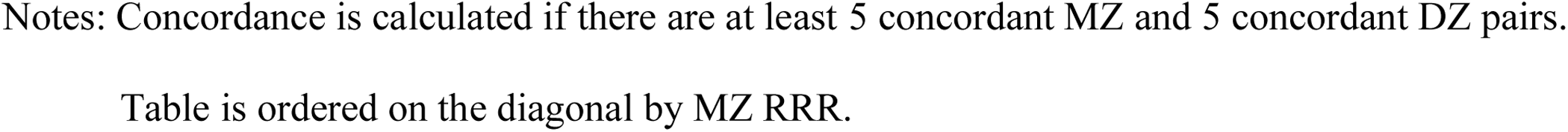
Lifetime relative recurrence risk of cross-cancer diagnosis among monozygotic (MZ), upper-right triangle and dizygotic (DZ), lower-left triangle, pairs of twins in the NorTwinCan Cohort, 1943 to 2016.

The generally higher RRRs among MZ pairs suggest that a substantial portion of the familial influences reflects genetic factors, under the assumption that MZ and DZ pairs share, on average, equal environmental exposures. Furthermore, several cancers--including breast, NHL, colon, prostate, bladder, lung, and skin non-melanoma--demonstrate multiple co-occurrence patterns with other cancer types, more so than other cancers. **Table S2** provides results from statistical tests assessing the equality of MZ and DZ concordance rates, corresponding to testing that the RRR don’t differ between MZ and DZ pairs, and evaluating the hypothesis that genetic effects are zero.

The familial effects contributing to the cross-cancer associations are decomposed into coheritable (**Table 5a**) and co-shared environmental (**Table 5b**) components. The magnitude of coheritable effects varies across cancer pairs. Prostate cancer exhibits some of the highest co-heritability with other cancers: 12% with rectum/anus cancer, 11% with kidney cancer, and 10% with multiple myeloma. Additionally, 10% of the variation in risk between colon and rectum/anus cancers, as well as between cervix uteri and skin melanoma cancers, is due to shared genetic factors. Several cross-cancer estimates yielded negative values, although these tend to be non-significant; they may be suggestive that the influence of genetic factors on cancer risk may operate in opposite directions for some cancer pairs: for example, pancreatic cancer showed inverse genetic effects with bladder cancer (−5.5%), cervix uteri cancer (−5.9%), and breast cancer (−5.6%).

**Table 5.**
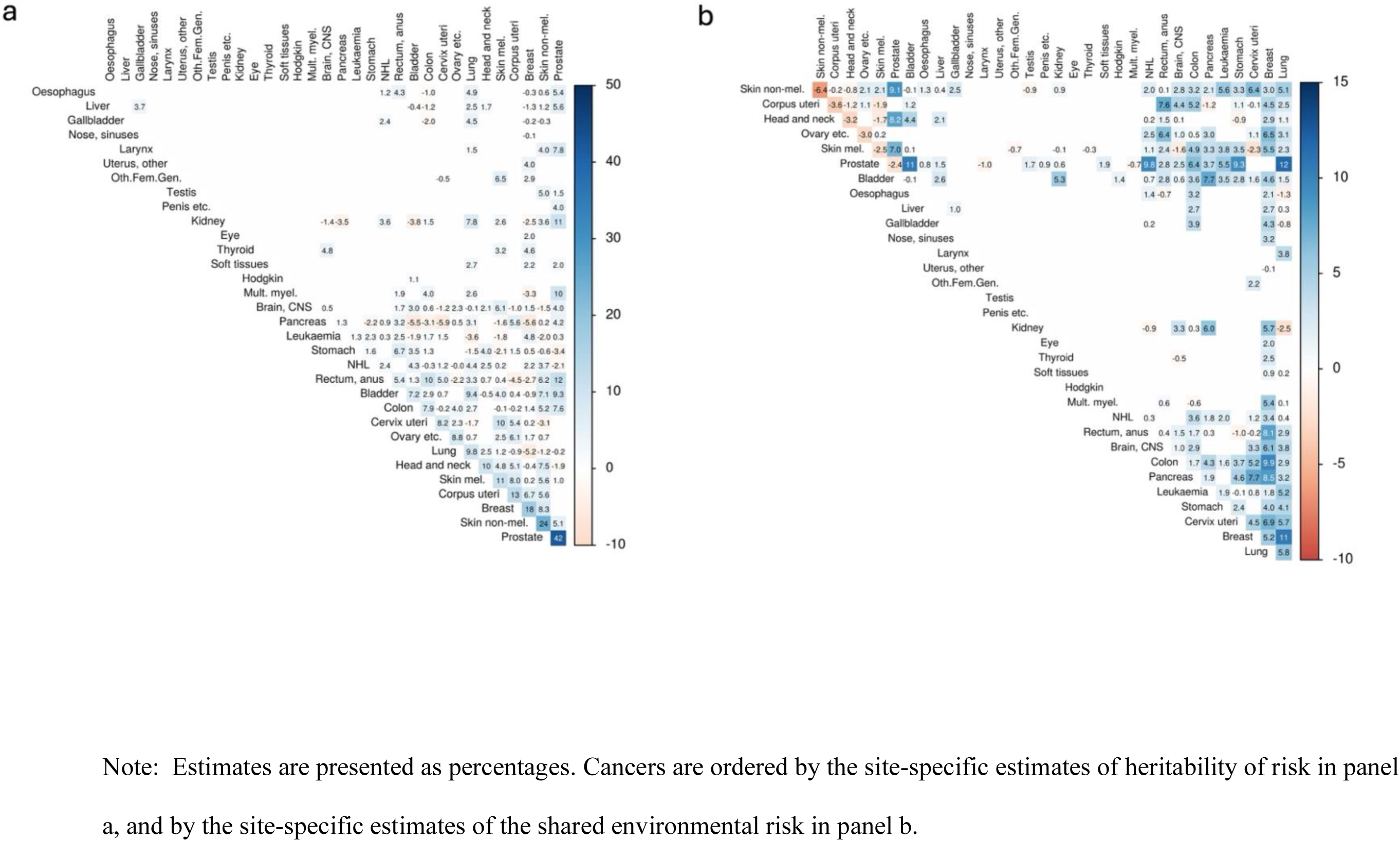
Coheritability (a) and co-shared environmental effects (b) on the risk scale across pairs of cancers in NorTwinCan, 1943 to 2016.

Co-shared environmental effects also contribute to associations between different types of cancers, and several cancers, including lung, prostate, and breast cancers, share environmental influences with multiple other malignancies. Among the largest effects, co-shared environmental factors accounted for 12% the variation in risk between prostate and lung cancer, 11% between prostate and bladder cancer, 9.8% between prostate and NHL, and 9.3% between prostate and stomach cancer. Similarly, co-shared environmental factors were estimated to explain 9.9% of the association between breast and colon cancer, 8.1% between breast and rectum/anus cancer, 11% between breast and lung cancer, and 8.5% between breast and pancreatic cancer. A few cross-cancer associations showed evidence of inverse effects from shared environmental factors, these were generally small, with the largest inverse effect observed between kidney and lung cancers (−2.5%). **Figure 1** presents circos plots illustrating the coheritability (panel 1a) and co-shared environmental relationships (panel 1b) across cancer types.

**Figure 1.**
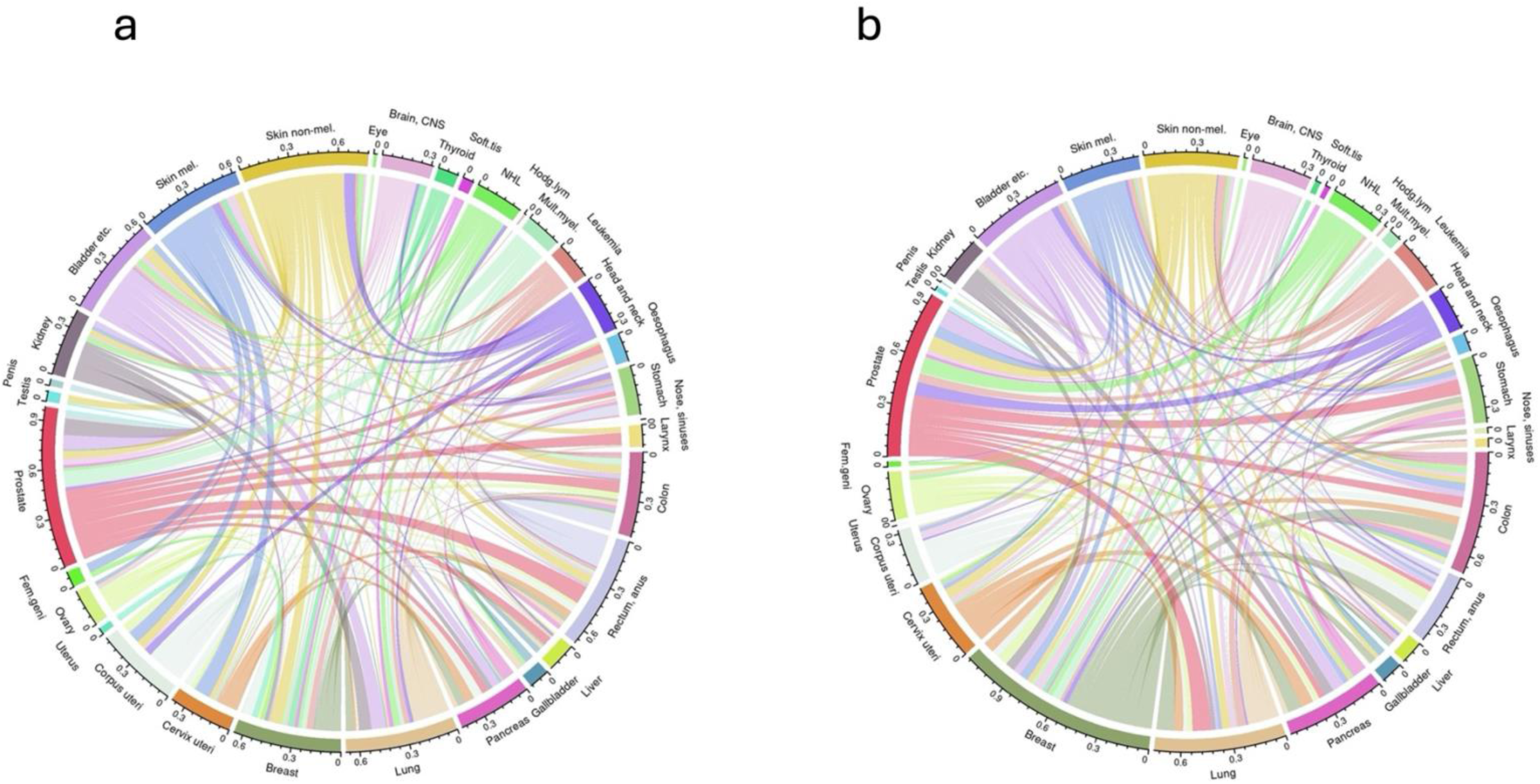
Circos plots showing cross-cancer co-heritabilities (a) and shared environmentability (b) calculated on the risk scale and corrected for censoring and competing risk of death. The width of the lines is proportional to the magnitude of shared genetic or environmental effects, NorTwinCan 1943 to 2016.

## Discussion

Our analyses of the NorTwinCan study^1,27^ provide refined estimates of familial cancer risk and offer novel insights into the importance of genetic and shared environmental factors underlying associations across 35 cancer sites. With data on 222,530 same-sex twins and 43,060 incident cancers followed for a median of over 41 years in four Nordic countries, this dataset supports a broad investigation into shared cancer risks, revealing both genetic and shared environmental contributions to cross-cancer associations. Consistent with earlier findings^1^, we observe significant site-specific discordance among cancer-concordant twin pairs. Heritable influences affect the risk of many cancers, coheritability across specific cancer pairs is widespread, and certain cancers share genetic influences with multiple other cancers. Shared environmental effects are also detectable for more cancers than previously reported^1^ and contribute to the shared risk across several sets of cancers. Familial risk was elevated for 23 out of the 35 cancer types as well as for cancer overall (**Table 2**). Among twins with a co-twin diagnosed with cancer, the cumulative incidence reached 51% among MZ pairs, and 45% in DZ pairs, compared to 37% for all individuals. Familial risk was consistently greater among MZ than DZ pairs, though confidence intervals overlapped for several cancer sites. The absolute increase in cancer risk relative to the general population was also somewhat greater among the MZ (15 percentage points) than the DZ (9 percentage points) twins.

The familial effects encompassed a broader range of cancers than in previous studie^1,2,29^ (**Tables 2 and 3**). Our findings indicate that shared family environment partially contributes to variation in cancer risk. In contrast, a study from the Utah population-based dataset attributed familial cancer risk predominantly to genetic mechanisms^30^. Unlike designs that focus exclusively on genotypic data, twin and family studies increasingly reveal shared familial effects as sample sizes expand. Previous evidence, notably from large Nordic studies, has highlighted shared family environment on cancer risk, particularly for cancers of the head and neck^1^, stomach^1,2,29^, lung^1,2,29,31^, breast^1,2,29,32^ cervix uteri^29^, corpus uteri^29^, ovary^29^, prostate^29^, bladder^2,29^, kidney^2^, thyroid^2^, nervous system^2^, endocrine glands^2^, testis^1^, rectal^1,2^, colon^1^ and colorectal cancer^29,33^, melanoma^2^, NHL^2^ and leukemia^2,29^.

Moreover, analyses of the Swedish Family-Cancer Database, which includes 9.6 million individuals, allowed for finer differentiation of shared familial effects, distinguishing between those attributable to shared (adult) familial and shared childhood environments for cancers of the stomach, colon, rectum, lung, breast, cervix, testis, kidney, bladder, nervous system, thyroid, endocrine glands, NHL and leukemia^2^. Our analyses also revealed shared environmental effects for overall cancer risk and for several cancers exhibiting a pattern of familial susceptibility. Cancers of the head/neck, esophagus, non-skin melanoma, ovary, kidney, bladder, and thyroid showed no significant shared environmental effects. The co-shared environmental variance we observed across cancer pairs may partly reflect lifestyle exposures, particularly obesity, alcohol use, and smoking, which rank among the most well-established risk factors for many of the cancers examined here. Previous analyses from the NorTwinCan cohort support a causal role for smoking in lung^31^ and other tobacco-related cancers^34^. Importantly, obesity, alcohol use and abuse, and smoking are themselves complex traits, influenced by numerous genetic variants and environmental factors. The cross-cancer genetic and shared environmental overlap we report may therefore reflect a combination of pleiotropy and shared liability to behaviors and exposures that influence multiple cancers through common risk-factor pathways.

Differences in heritability estimates across models illustrate important considerations when comparing across studies. Heritability estimates derived from the parametric approach are consistently higher and exhibit wider confidence intervals, even when tetrachoric correlations from the liability model are used to calculate the heritability of risk. However, once estimates are transformed to a uniform risk scale, they converge considerably. Notably, after accounting for the competing risk of death, heritability estimates from the non-parametric model align more closely with those from the Swedish population^2^ and genotype-based analyses^3–6^. Failure to account for this competing risk can introduce upward or downward bias in heritability estimates.

Heritability values from the twin parametric model generally exceed those derived from family^2^ and genotype-based^3–6^ data. This finding is consistent with the well-documented “hidden heritability” gap observed between estimates from twin studies versus those derived from identified common genetic variants^35–37^. The discrepancy may reflect the broader scope of genetic variation captured by twin studies, which includes both additive and non-additive effects, as well as additive effects arising from variants not captured by genotyping arrays. Moreover, GWA-based heritability estimates may also be limited by the underrepresentation of rare variants. Including these considerably increases heritability estimates for common traits and diseases^38^ and enhances the performance of polygenic risk scores for complex traits beyond those based solely on common variants^36,39,40^.

Our analyses of the familial factors underlying the relationships between 35 malignancies represent the largest number of cancers analyzed in a single study. We find familial effects, comprising coheritable and co-shared environmental factors, are widespread across many cross-cancer associations. Several cancers exhibit relationships to multiple other malignancies, particularly those with higher heritabilities.

Most genetic studies examining pleiotropy across cancers have focused on a limited number of cancer sites within a single study^3–5,10,11^. Comparing results between these genetic studies is challenging due to differences in the cancers studied, sampling designs, and methods used to calculate genetic associations. A recent, large-scale analysis of genome-wide association data for twelve solid cancers revealed widespread but modest shared genetic influences. Genetic correlations greater than 0.4 were reported for five cancer pairs, with lung and renal cancers showing genetic links to multiple other cancers, and genetic correlations varied between other cancers and subtypes of breast and lung cancer. Overall, heritable factors were primarily cancer and tissue-specific^12^. Cross-cancer analyses of 16 cancer-specific PRS scores identified 11 cancer pairs that exhibited significant pleiotropy, with the oral cavity/pharynx-lung pair showing a negative direction of effects^8^.

Despite the limitations in comparing GWAs-based pleiotropy studies, some consistent patterns have emerged, such as the shared genetic basis for specific cancer pairs, including colorectal-lung^3,6,10,12^, breast-colorectal^3,12^, breast-lung^3,6,9,12^, breast-ovarian^3,9,12^, colorectal-pancreatic^6,10,12^, prostate-breast, and prostate-lung^19^. However, in contrast to our findings, prostate cancer did not exhibit widespread genetic correlations with other cancers.

Beyond the pairwise associations between cancers, and consistent with other studies, we find that certain cancers, including colorectal, lung, prostate and breast cancers, are genetically linked to a broader range of other cancers^5,6,12^, demonstrating more extensive pleiotropy. While beyond the scope of our data, molecular studies have begun to elucidate pleiotropic risk variants, including loci associated with the susceptibility to develop breast, colorectal, ovarian and prostate cancers^41^, as well as genomic regions and potential pathways implicated in susceptibility to multiple cancer types^8,10,12^.

Shared environmental influences were evident across several cross-cancer associations. Although environmental risk factors are well established in cancer etiology, few family or twin-based studies have assessed their shared contributions across cancers. Our findings underscore the importance of incorporating familial environmental factors in cross-cancer investigations.

This study has several notable strengths. The extended follow-up is a key strength that enabled near-complete ascertainment of cancer outcomes over several decades. The large sample size provided a robust basis for assessing incident cancer cases and avoided potential biases that can arise when incident and prevalent cases are combined, as is common in genome-wide association studies. The target trial emulation is less prone to confounder effects. The population representative data span a broad spectrum of cancers, including all common cancers, and analyses were adjusted for the competing risk of death. These strengths enabled us to derive new or updated estimates of heritability and shared environmental effects for multiple cancers, as well as for cross-cancer associations.

The twin design uniquely enables the disentanglement of genetic and shared environmental effects and is less prone to confounding of these components than family-based studies. Unlike molecular genetic studies, twin analyses provide a more comprehensive picture of familial risk by jointly estimating genetic and shared environmental effects—an aspect not captured by GWAs data, but for which our findings indicate a measurable impact on cancer risk. Analyses of whole genome-wide data suggest that the gap between twin-based and molecular genetic estimates of the genetic contributions is decreasing^38^, although this newest study did not examine cancers.

A potential limitation is that temporal changes in shared environmental exposures that affect cancer risk (e.g. smoking) are not captured by our estimates, which reflect time-averaged rather than time-specific effects. The study is further limited to individuals of Northern European ancestry, which, while reducing genetic and cultural heterogeneity, limits the generalizability of our findings to more diverse populations. Furthermore, the exclusion of opposite-sex twin pairs restricted our ability to investigate sex-specific genetic and environmental effects on cancer.

Our findings illustrate a broad interconnectedness of cancer risks, partly driven by familial factors. Genetic and shared environmental influences affect the risk of developing individual cancers and contribute to cross-cancer associations. These results highlight the importance of incorporating measures of shared familial influences into cancer risk models, and of moving beyond single-cancer approaches to better capture the complexity of risk patterns spanning multiple cancer types.

Future research should focus on identifying the specific genetic and shared environmental influences underlying these relationships to further elucidate cancer etiology, refine risk prediction, and inform targeted prevention strategies.

## METHODS

### Study Population

The NorTwinCan collaboration was established to explore how genetic and shared familial influences affect variation in cancer risk^1,27,42,43^. The data were compiled by linking information from the national twin registries in Denmark, Finland, Norway, and Sweden to population-based data from the national, comprehensive cancer registries. The cancer registries are continually updated and renewed linkages to the twin registries enable virtually complete long-term follow-up of the twins for cancer incidence and mortality. Linkages and relevant ethics approvals have been granted by the relevant data protection authorities and ethics committees in each country and fully comply with the General Data Protection Regulation of the European Union (EU-GDPR, 2016).

Details of the process for compiling the twin registries are described elsewhere^1,44^. The twin registries differ regarding the span of birth cohorts included and the date at first and last follow-up (Table 1). Zygosity was originally established through questionnaire methodology, which has shown high validity with DNA-based zygosity classifications, and typically correctly classifies zygosity for more than 95% of the twins^45–47^.

The full NorTwinCan cohort includes nearly 400,000 individuals from twin or triplet births. Among these, zygosity is known for 316,397 twins. We restricted this study to MZ and same-sex DZ (70 percent of the DZ twins) pairs, resulting in a cohort of 222,530 twins.

### Follow-up and measures of cancer

The most recent linkages with the cancer registry were performed in 2018^27^ and included all cancer registrations through 2016 for Denmark and Finland and through 2015 for Norway and Sweden (**Table 1**). The twins were followed up to the end of the study using registry data that provided information about cancer diagnosis, death, or emigration between 1943 and 2016.

The nearly complete cancer registrations^44^ provide diagnoses classified according to the International Classification of Diseases, 10^th^ Revision (ICD-10). To overcome differences in coding practices between the contributing Nordic countries, the NORDCAN system^48,49^ was used to classify the ICD codes according to cancer sites, enabling cross-country comparisons. We estimated the risk of any cancer (overall cancer) and cancers classified into 35 anatomical sites.

### Target trial emulation

To emulate a target randomized trial and enhance robustness to confounding, we exploit the natural structure of the twin cohorts, which are representative of the background population^44^. Co-twins can be regarded as quasi-randomly assigned to follow-up at different calendar times, (e.g. at birth, inclusion in, or at inception of the twin registry). Consequently, co-twins contribute equal risk time at baseline, while outcomes are observed prospectively over an extended follow-up period, spanning decades.

Estimation of the risk measures is based on biometric models adapted for time-to-event twin data. These models exploit variation in genetic relatedness and shared environments within twin pairs by comparing the risk of diagnosis between twins who are more versus less similar in their genetic and environmental influences. This approach enables estimation of risk while accounting for shared familial factors.

### Statistical Analyses

We conducted two complementary sets of analyses. The first focused on site-specific cancers and estimated cumulative incidences, familial risk, heritability, shared and non-shared environmental contributions for each site. The second evaluated cross-cancer associations, estimating familial effects across pairs of cancers among 35 sites, and provided estimates of co-heritable and co-shared environmental contributions to cross-cancer risk.

### Site specific analyses

#### Cumulative incidence and familial risk of cancer

The lifetime risk of a cancer diagnosis before age 100 years stratified by sex, zygosity, and country was estimated from the cumulative incidence function by age, using non-parametric counting process modelling^50^. These analyses account for delayed entry due to variable initiation dates across countries, right-censoring caused by the end of follow-up or emigration, and competing risk of death. In the case of site-specific cancers, the familial or casewise concordance risk reflects the risk to a twin of developing cancer at that site, given that their co-twin has been diagnosed with the same cancer. Excess familial risk of cancer is estimated by comparing these conditional risks with the cumulative incidence estimated from the twin data considered as individuals. The incidence rates among the twins reflect the incidence in the background population of the respective countries^44^. Stratification of the conditional risks by zygosity provides insight into the nature of the familial factors—genetic and shared environment, that affect cancer risk. The genetic relationship between members of a DZ pair is the same as with full siblings. Hence, the familial risk among DZ twins also represents the sibling risk.

### Heritability estimates

Estimates of genetic influences underlying disease risk vary by analytic method and the scale used to assess risk^51^. To facilitate comparisons with previous studies, we estimated site-specific cancer heritabilities using both non-parametric (risk scale) and parametric (liability scale) approaches (see **Statistical Supplement** for details). Both approaches account for right censoring after the end of follow-up due to staggered registry initiation, variable follow-up time, and loss to follow-up. Estimates based on the non-parametric approach additionally adjust for the competing risk of death.

*The non-parametric approach* for time-to-event data estimates heritability and shared environmental contributions to cancer risk directly on the risk scale, accounting for censoring and competing risk of death^28^. Estimates were derived from Equation 1 (heritability) and Equation 2 (shared environment) (**Statistical Supplement**) and applied to cancers with at least five concordant MZ and DZ pairs.

*The parametric approach* uses the classic liability-threshold model, which assumes an underlying, normally distributed cancer liability determined by multiple genetic and environmental factors^52^. Biometric modeling employed a bivariate biprobit model with inverse probability weighting, IPCW^53^ to adjust for censoring. Unlike the non-parametric model, which estimates variation directly on the risk scale, the liability-threshold model constrains the total variance in liability to one. This variance is decomposed into additive genetic (A), dominant genetic (D), shared environmental (C), and non-shared environmental (E) components. Heritability of liability represents the proportion of total variance explained by the genetic components, while the shared environmental component reflects exposures common to both twins.

### Cross cancer-analyses

#### Measures of cross-cancer familial dependence

We extended the site-specific risk models (**Statistical Supplement**) to investigate cross-cancer familial dependence using the non-parametric risk-scale approach^50^ and estimates of casewise concordance and relative recurrence risks. The *cross-cancer concordance* reflects the probability that a twin pair develops a specific combination of cancers before a given age^50^, while the *relative recurrence risk* (RRR) compares the probability of these cancers occurring to that expected under independence.

Differences between MZ and DZ pairs in the measures of cross-cancer familial dependence provide insight into the nature and magnitude of the familial influences underlying cross-cancer associations. Familial factors are implicated when the risk among DZ twins exceeds the cumulative incidence or when the RRR exceeds one. Stronger dependence in MZ than DZ twins suggests that genetic factors contribute importantly to the cross-cancer association^28,54^. Co-heritability and co-shared environmental contributions across cancer pairs were obtained by extending the non-parametric analyses to a bivariate model. Hypothesis testing and estimation of uncertainty were conducted using standard procedures^50,54^. Model sensitivity was assessed by comparing estimates generated under the various models, with and without adjustment for competing risk of death (**Statistical Supplement**).

Sensitivity analyses (**Statistical Supplement**) compared heritability estimates derived from non-parametric risk-scale models with those from parametric liability-threshold models that account for censoring (**Figure S1**). We also evaluated bias from ignoring the competing risk of death across cancer sites (**Figure S2**).

## Acknowledgements

This work was supported by grants from the Nordic Cancer Union (PI: J.R. Harris); *Genetic Epidemiology and Familial Risk of Cross-Cancer Associations: A Nordic Twin Study*; and the Research Council of Norway through its Centres of Excellence funding scheme, project number **262700**.

## Data Availability

Additional data access requests can be made directly to the national cancer registers and twin cohorts that are responsible for data administration and sharing.

## Author Contributions

JRH, JvBH, and JK conceived the study and designed the research. JvBH supervised the methodological work. JvBH and SBC performed the statistical analysis and generated the figures. JvBH, SBC, JRH and JK interpreted findings. JRH, JvBH, JK and SBC drafted the manuscript. All authors critically reviewed the manuscript for intellectual content, contributed to the manuscript revision, and read and approved the final version.

