## Supplemental information, tables and figures for "Genetic and Shared Environmental Influences on Cancer Risk and Cross-Cancer Associations in Nordic Twins"

### Statistical Supplement

#### Biometric modeling

##### *Site Specific Analyses*

**Non-parametric approach.**

Heritability of risk ($h^2$) adjusted for the competing risk of death was calculated as the ratio of twice the difference in concordance risk between MZ and DZ pairs divided by the total variance in risk[1-3] (Equation 1). This derivation is provided elsewhere[1,2]. Essentially, it projects the biometric ACE model onto the risk scale and therefore assumes that the ACE model is applicable at each time point. Shared environmental contribution ($c^2$) was estimated in the same way as twice the difference in concordance risk in DZ pairs minus the concordance risk in MZ pairs, divided by the total variance in risk (Equation 2). These equations use time-to-event concordance risks at time $t$, and the cumulative cancer incidence $F(t)$, and assume equal cumulative cancer incidences in MZ and DZ twins:

$$h^2(t) = \frac{2(C_{MZ}(t) - C_{DZ}(t))}{F(t)(1 - F(t))} \text{(Equation 1)}$$

$$c^2(t) = \frac{2C_{DZ}(t) - C_{MZ}(t)}{F(t)(1 - F(t))} \quad \text{(Equation 2)}$$

**Parametric approach.**

The classic liability-threshold model assumes equal cancer incidences (thresholds or prevalences) in MZ and DZ twins, that these prevalences are site-specific, and can vary by sex, and cohort[4]. The biometric IPCW liability-threshold model specifies how the differential degree of genetic relatedness by zygosity (MZ or DZ) contributes to differences in twin covariances for cancer between MZ and DZ pairs compared to the total variance of the cancer risk. This model also generates tetrachoric correlations and casewise concordances for MZ and DZ pairs which can be utilized to calculate heritability on the risk scale as described for the non-parametric approach.

##### *Cross-Cancer Analyses*

Cross-cancer relationships were estimated using a non-parametric risk scale approach based on casewise concordance and RRR. The RRR was defined as the ratio of the cross-cancer concordance to the product of cumulative incidences for cancers A and B. Lifetime RRRs reflect the cumulative incidence at age 100 and were estimated across all cross-cancer pairs by zygosity. Values are calculated only when there are at least five cross-cancer concordant MZ and five cross-cancer concordant DZ pairs.

If an individual twin had multiple cancer diagnoses, then all combinations within the pair were included in the cross-cancer analyses.

All analyses were performed using the statistical software R version 4.0.1[5] with the mets package version 1.3.2[1,4].

##### ***Sensitivity analyses***

Sensitivity analysis assessed the robustness of the heritability estimates to modeling assumptions by comparing estimates from the parametric liability-threshold framework and those from the non-parametric risk-scale approach. Both methods accounted for right censoring, while the non-parametric approach additionally adjusted for the competing risk of death.

After projecting liability model estimates onto the cumulative risk scale, heritability estimates from the two approaches converged closely (**Figure S1**), indicating that the observed differences largely reflect scale rather than substantive differences between models[6].

Next, we evaluated bias introduced from ignoring the competing risk of death by comparing estimates from the non-parametric approach with and without adjustment for death. Failure to account for competing mortality revealed bias in the heritability estimates, with both the direction and magnitude of bias varying across cancer sites (**Figure S2**).

Overall, these analyses indicate that parametric and non-parametric approaches yield consistent heritability estimates when evaluated on a common scale, and that accounting for competing mortality is important for unbiased estimation of genetic and environmental contributions to cancer risk.

**Table S1.** Number of cancers within pairs among twin 1 vs. twin 2, NorTwinCan 1943 to 2016.

| MZ | 0 | 1 | 2 | 3 | 4 | Total |
|---|---|---|---|---|---|---|
| 0 | 30,813 | 4,251 | 380 | 31 | 2 | 35,477 |
| 1 | 3,437 | 1,781 | 262 | 22 | 4 | 5,506 |
| 2 | 353 | 305 | 72 | 9 | 0 | 739 |
| 3 | 31 | 28 | 9 | 3 | 1 | 72 |
| 4 | 0 | 3 | 1 | 1 | 0 | 5 |
| 5 | 0 | 1 | 0 | 0 | 0 | 1 |
| Total | 34,634 | 6,369 | 724 | 66 | 7 | 41,800 |

| DZ | 0 | 1 | 2 | 3 | 4 | Total |
|---|---|---|---|---|---|---|
| 0 | 43,153 | 7,827 | 789 | 75 | 7 | 51,851 |
| 1 | 6,090 | 2,670 | 356 | 37 | 1 | 9,154 |
| 2 | 731 | 371 | 66 | 8 | 1 | 1,177 |
| 3 | 58 | 32 | 8 | 4 | 0 | 102 |
| 4 | 7 | 4 | 1 | 0 | 0 | 12 |
| 5 | 0 | 1 | 0 | 0 | 0 | 1 |
| Total | 50,039 | 10,905 | 1,220 | 124 | 9 | 62,297 |

**Table S2.** P values for tests of equality of MZ and DZ concordance. P-values provided as percentages.

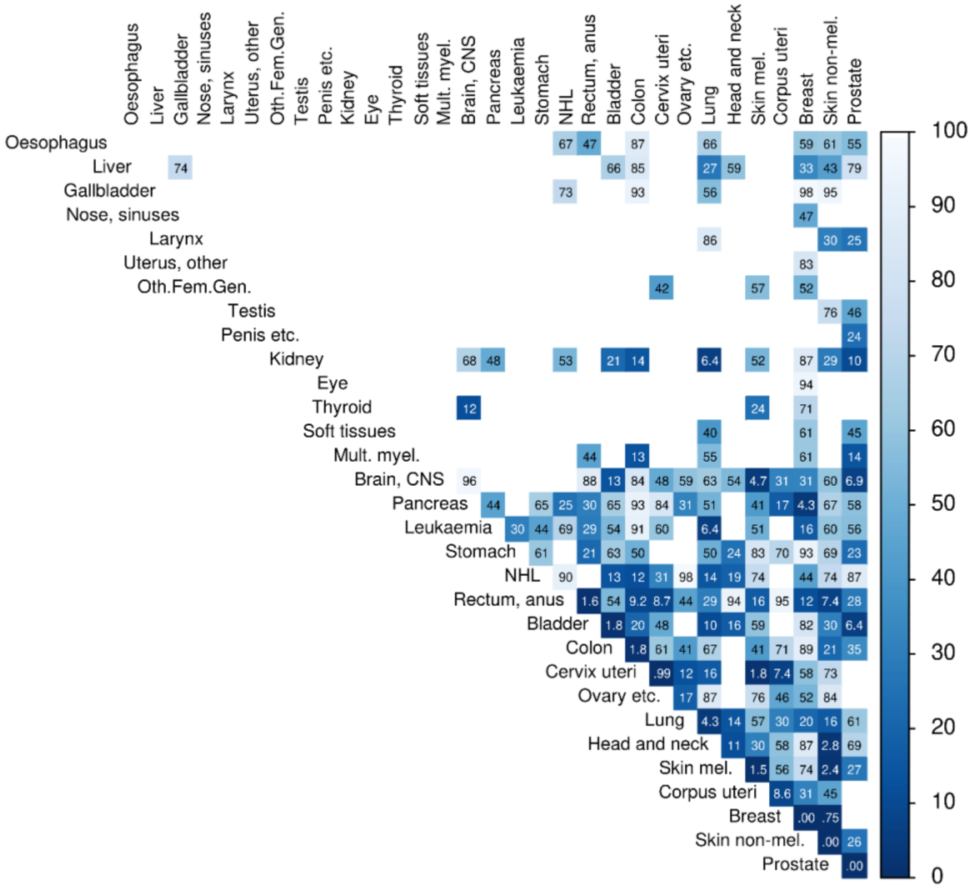


Oth.Fem.Gen, other female genital; Mult. Myel, multiple myeloma; mel., melanoma, CNS, central nervous system; NHL, non Hodgkin lymphoma

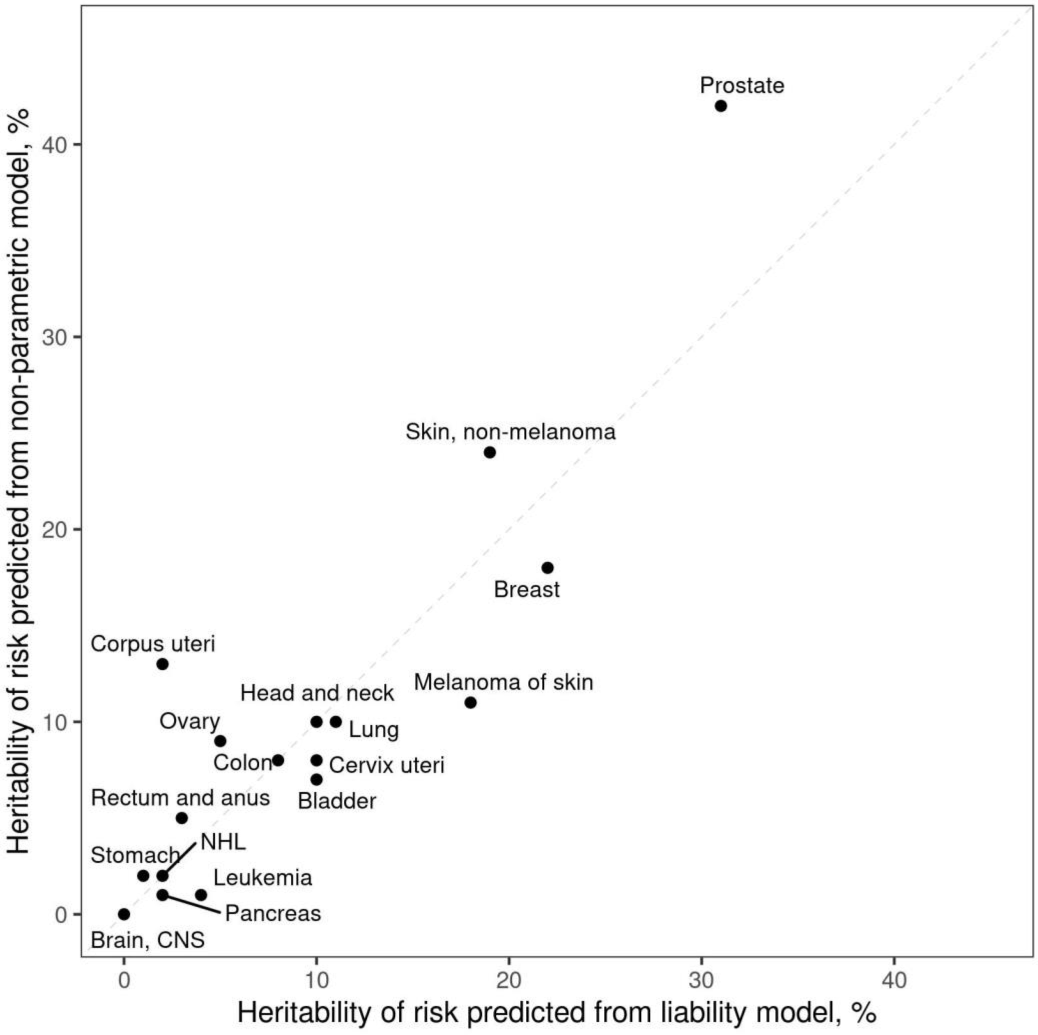


**Figure S1**. Convergence of heritability estimates from non-parametric and liability models when all estimates are projected on the risk scale, NorTwinCan 1943-2016.

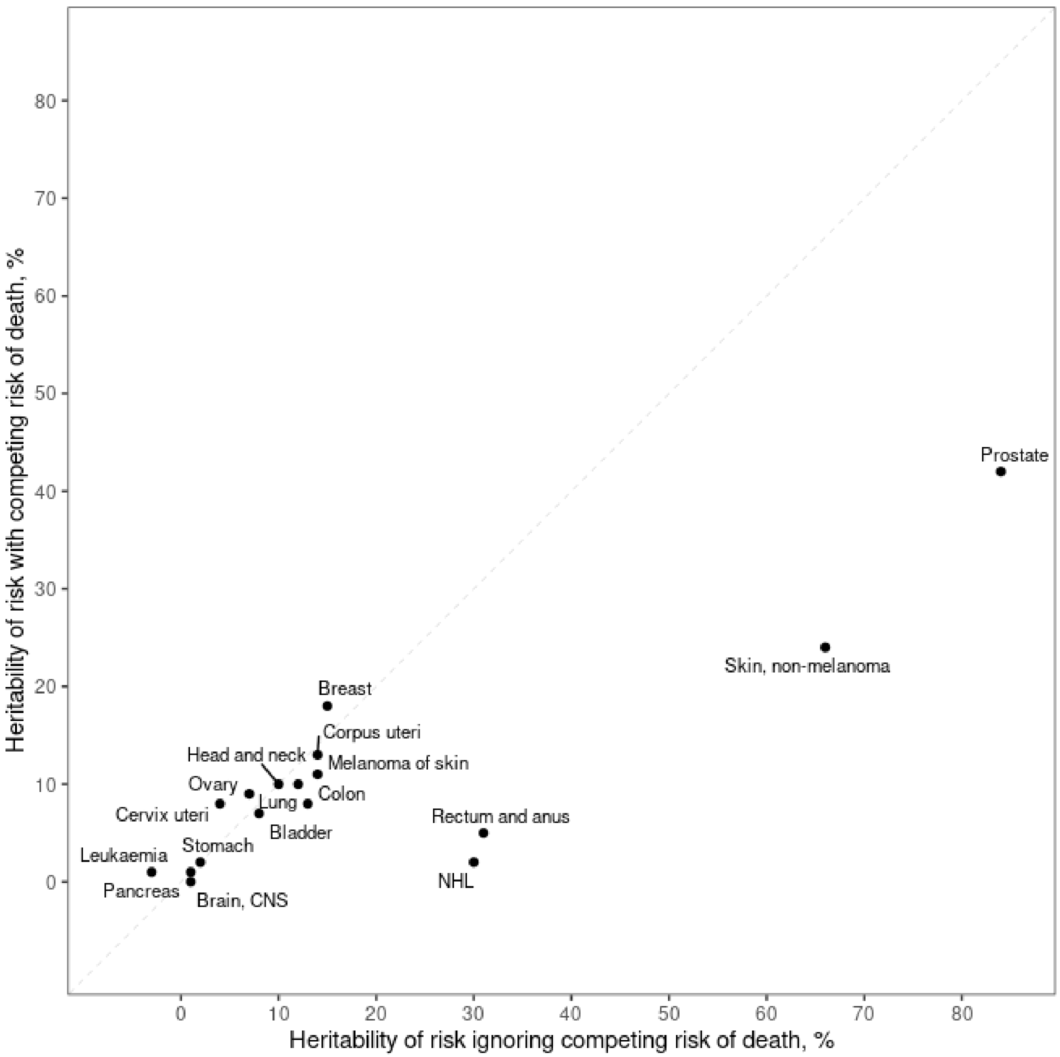


**Figure S2**. Bias in estimates of the heritability of cancer risks when the competing risk of death is not modeled.
